# Pain-indicated buprenorphine prescribing in the United States, 2017–2025: national trends, state policy approaches, and payer patterns in a longitudinal ecological study

**DOI:** 10.64898/2026.11.12.26367897

**Authors:** Meredith C. B. Adams, David M. Kline, Jae C. Yong, Cara McDonnell, Robert W. Hurley

## Abstract

**Background:** Buprenorphine offers an analgesic option for selected adults with chronic pain. National trends in pain-indicated buprenorphine prescribing, variation across state policy approaches, and consistency within multiple insurance payers have not been examined together. We described national and state prescribing trends, compared growth among states with documented policies, and examined whether state patterns persisted among Medicaid recipients.

**Methods:** Longitudinal ecological analysis of 2017-2025 prescription claims for 867,591 unique adults receiving transdermal or buccal buprenorphine across 888 three-digit ZIP code areas, with a systematic policy review of the 50 states and District of Columbia. Outcomes were annual recipients of pain-indicated formulations per 100,000 adults, observed and modeled with a hierarchical Bayesian model (95% credible intervals), state policy characteristics, and payer-restricted rates.

**Results:** The observed national rate increased from 53.59 to 78.04 recipients per 100,000 adults (45.6%; 4.81% annualized); model-derived average annual growth was 5.04% (95% credible interval, 2.88%-7.28%). The policy review identified distinct guidance, coverage, oversight, and educational approaches. Annualized growth was 25.43% in Minnesota, 13.50% in Colorado, 6.29% in North Carolina, 3.35% in Virginia, 2.99% in California, 0.63% in West Virginia, 0.50% in Oregon, and −2.96% in Mississippi.

Identifiable-Medicaid prescribing grew 3.10% annualized nationally versus 4.81% for all payers and increased in Colorado and Minnesota, but declined in Virginia and North Carolina, where plan coding and managed-care transitions limited Medicaid identification.

**Conclusions:** Prescribing increased nationally but varied substantially across states with different policy approaches, and Medicaid-restricted patterns did not consistently mirror all-payer trends. Policy content, timing, and payer reach warrant evaluation; however, these ecological comparisons do not establish causation.

## INTRODUCTION

Chronic pain is common in the United States, and clinicians must balance symptom relief, function, and medication-related harm [1]. For selected adults, buprenorphine offers analgesia with lower respiratory-depression risk than full-agonist opioids, although it remains an opioid with important risks [2–4]. Understanding who receives pain-indicated formulations, and how prescribing varies geographically, can identify questions about access that aggregate opioid statistics do not resolve.

The national context for opioid prescribing changed during the past decade. The 2016 CDC guideline favored nonopioid therapy for chronic pain, and the 2022 update discussed transition to buprenorphine for selected patients [5, 6]. How national guidance reaches practice depends partly on state medical boards, health departments, and insurers, which may influence prescribing through clinical guidance, coverage, prescribing rules, and education [7, 8]. Much buprenorphine policy research has focused on opioid use disorder (OUD), whereas pain-indicated formulations carry different costs and access requirements [9–12]. A recent Medicaid study examined 2015-2024 prescription trends using coverage restrictions classified in August 2025 [13]. Less is known about how all-payer prescribing has changed nationally, how growth varies among states with different policy content and timing, and whether state patterns hold within Medicaid, the payer that many state policies directly reach.

We therefore addressed three linked questions using national prescription claims and a policy review of all 50 states and the District of Columbia. First, we described national trends and geographic variation in pain-indicated buprenorphine prescribing from 2017 through 2025, hypothesizing that prescribing would increase but that growth would vary across US regions and states. Second, we characterized state approaches to clinical guidance, coverage, prescribing oversight, and education, and compared observed growth among states with documented policies. Third, because many of these policies govern Medicaid coverage or oversight, we examined whether all-payer state patterns persisted among identifiable Medicaid recipients.

## METHODS

### Study Design and Oversight

We conducted a longitudinal ecological analysis of 2017-2025 prescription claims and a policy review covering all 50 states and the District of Columbia. The analysis proceeded in three stages: national and geographic prescribing trends, state policy review with state-level comparisons, and payer-restricted analyses. Policy review used the READ framework [14].

Reporting was guided by STROBE (Table A in S1 Appendix) [15]. The Wake Forest University School of Medicine Institutional Review Board (IRB00127502) determined that the study, which used deidentified prescription claims, was exempt from review and waived the requirement for informed consent.

### Prescription Data and Measures

We used the IQVIA Longitudinal Prescription Claims (LRx) database, which contains deidentified prescription records with patient age and three-digit ZIP code (ZIP3). Data was first accessed on September 21^st^, 2023 and last accessed on September 6^th^, 2026. IQVIA reports that LRx captures approximately 94% of retail, 82% of mail-service, and 74% of long-term-care prescription claims; coverage varies by channel and over time [16, 17]. We identified transdermal patches and buccal films by National Drug Code and verified their FDA-approved indications (Table C in S1 Appendix). Throughout the study, “pain-indicated” describes a formulation approved for pain, not a diagnosis confirmed in the claims; off-label use of OUD formulations for pain was outside the study definition.

We included adults aged 18 years or older and removed duplicate recipients within each year. The outcome was the annual number of unique recipients per 100,000 adults. The hierarchical model used fixed 2022 American Community Survey (ACS) five-year adult population estimates aggregated to ZIP3 areas; a sensitivity model used fixed 2017 estimates [18]. The separate observed-rate analyses used year-matched ACS five-year releases for 2017-2024 and the 2024 denominator for 2025. Each ACS five-year release summarizes a rolling five-year period, not a single-year population survey. The authors had no access to information that could identify individual participants during or after data collection.

### National Prescribing Trends

We analyzed annual recipient counts using a hierarchical Bayesian Poisson model that allowed starting rates and growth to differ among ZIP3 areas while sharing information within states and nationally [19]. The state-level hierarchy included Medicaid expansion status for intercepts and slopes. The model assumed constant proportional change within each ZIP3 area. We converted fitted log-slopes to annual percentage growth with 95% credible intervals (CrIs). The model-based national summary used the unweighted arithmetic mean of fitted intercept and slope coefficients across the analytic geographic areas. It includes jurisdictions with and without explicit recognition and is distinct from the observed population-weighted national rate. Model specifications and diagnostics appear in the S1 Appendix.

Observed national and state rates used annual unique recipients and year-matched adult populations. Because ZIP3 areas can cross state boundaries, Census ZCTA-county relationships and residential shares from the June 2026 HUD-USPS crosswalk informed ZIP3-to-state allocation, with year-specific ACS adult-population weights [18, 20]. A recipient with multiple valid ZIP3 records contributed one total person after normalizing state weights. The national comparator divided unique mapped recipients by the combined allocated adult population of the 50 states and DC. Sensitivity analyses held 2024 populations constant or assigned each recipient to the state with the largest cumulative geographic weight. Tables J-M in S1 Appendix report rates, rankings, sensitivity results, and completeness.

### State Policy Review

Two investigators (M.C.B.A. and R.W.H.) reviewed medical board policies, state-issued or state-endorsed guidelines, Medicaid pharmacy documents, and legislation across all 51 jurisdictions.

They independently adjudicated policy evidence using an evidence-level decision log; a third investigator (C.M.) was designated to resolve disagreements. Explicit clinical recognition required a recommendation or preference for buprenorphine as an analgesic. Institutional guidance without state endorsement, unenacted proposals, and workers’ compensation-only policies were outside the policy scope (Table B in S1 Appendix).

We characterized clinical guidance, coverage, prescribing oversight, and education. The review focused on 2014-2023, retaining earlier provisions and updates through 2025. We distinguished document publication, legal effective dates, and clinical uptake. Routine formulary placement was recorded as coverage, even when it did not meet the rubric for enhanced access. Policies with different payer reach or clinical purposes were not combined into a single treatment exposure (Tables B, D, E, and I in S1 Appendix).

From the nationwide review we selected eight states to illustrate the range of documented approaches and their timing relative to follow-up. Colorado illustrated clinical guidance, coverage, and education; Minnesota and California illustrated clinical guidance, prescribing oversight, and education; and Oregon illustrated clinical guidance and education. Virginia illustrated clinical guidance and coverage policies. Mississippi and West Virginia illustrated Medicaid formulary approaches that originated before follow-up, and North Carolina illustrated routine coverage without an enhanced pain-specific pathway. These states are examples drawn from the full review, not a representative sample of jurisdictions.

### State Comparisons and Timing Analyses

We ranked all 51 jurisdictions by annualized observed growth and compared the illustrative states with the national comparator. We also fitted additional Bayesian log-rate trends to the 2017-2025 observed series for each jurisdiction and the nation, using year-varying denominators and autoregressive residuals; these models supplement the hierarchical ZIP3 model (Table N in S1 Appendix). For dated policy actions, we compared the last full pre-event year with the first full post-event year and the corresponding change in the pooled remaining jurisdictions. We omitted partial implementation years and used a second comparator excluding 15 policy examples or candidates (Table H in S1 Appendix).

In additional analyses, we modeled the log ratio of Colorado to rest-of-US rates with seasonal terms and autoregressive errors and examined nominated breaks in January 2018 and January 2023. Sensitivities used alternative break dates around documented changes and alternative model assumptions (Table G in S1 Appendix). Annual segmented comparisons addressed Colorado, California, and Oregon when at least three full pre-and post-event years were available (Table Q in S1 Appendix). Consistent with other opioid-policy evaluations, we distinguished implementation dates and examined comparison trajectories [48, 49]; we did not pool heterogeneous guidance, coverage, and educational actions into a single staggered intervention.

### Payer-Restricted Analyses

Because many state policies apply specifically to Medicaid, all-payer rates could dilute associations concentrated among Medicaid recipients or, conversely, could rise while prescribing for the targeted payer changed little. We therefore examined whether all-payer patterns persisted among Medicaid recipients. Payer-restricted analyses covered 2017-2025, using patient and plan dictionaries for the corresponding records. Identifiable Medicaid comprised fee-for-service and explicitly identified managed Medicaid plans; a narrower analysis used fee-for-service only.

Mixed Medicaid/Medicare categories were not automatically classified as Medicaid. Rates used adult population, not Medicaid enrollment, as the denominator, so payer-restricted rates describe recipients per adult resident rather than per enrollee (Tables O and P in S1 Appendix).

### Use of artificial intelligence tools

Generative artificial intelligence tools were used during manuscript preparation as follows. Claude assisted with editorial revision of text (spelling and grammar), initial web searching for state policy sources, and initial figure and table preparation. Policy sources identified with AI assistance were retrieved and independently supplemented and adjudicated against the original documents by M.C.B.A. and R.W.H. AI tools did not generate or alter any data, model estimates, or values shown in figures or tables. All AI-edited output was reviewed and confirmed by the authors, who take responsibility for all hypotheses, interpretations, results, conclusions, and limitations, for the accuracy of all statements, and for the citation of all sources.

## RESULTS

### National Prescribing Trends

The model sample included 867,591 unique adults, deduplicated within each year across 2017-2025, in 888 ZIP3 areas; annual eligible adult recipients ranged from 136,241 in 2017 to 207,119 in 2025 (Table M in S1 Appendix). The observed population-weighted national rate increased from 53.59 in 2017 to 78.04 in 2025 per 100,000 adults, a 45.6% increase (4.81% annualized).

The model-derived geographic average annual growth was 5.04% (95% CrI, 2.88%-7.28%), converted from the unweighted mean fitted log-slope. The observed comparator describes the pooled population; the model summary describes average fitted geographic growth. Annual observations showed departures from constant growth. The national rate changed little in 2017-2018 and 2021-2022, then increased from 69.82 in 2022 to 78.04 in 2025 (Table J in S1 Appendix).

Geographic variation was substantial (Fig 1). In 2017, estimated rates were generally higher in Southern and South-Central areas and lower in much of the Northeast and West Coast. Growth was pronounced in parts of the Upper Midwest and South, with slower growth in portions of the Northeast, Pacific Northwest, and Appalachia. These within-region differences caution against treating regional averages as uniform experiences.

**Fig 1.**
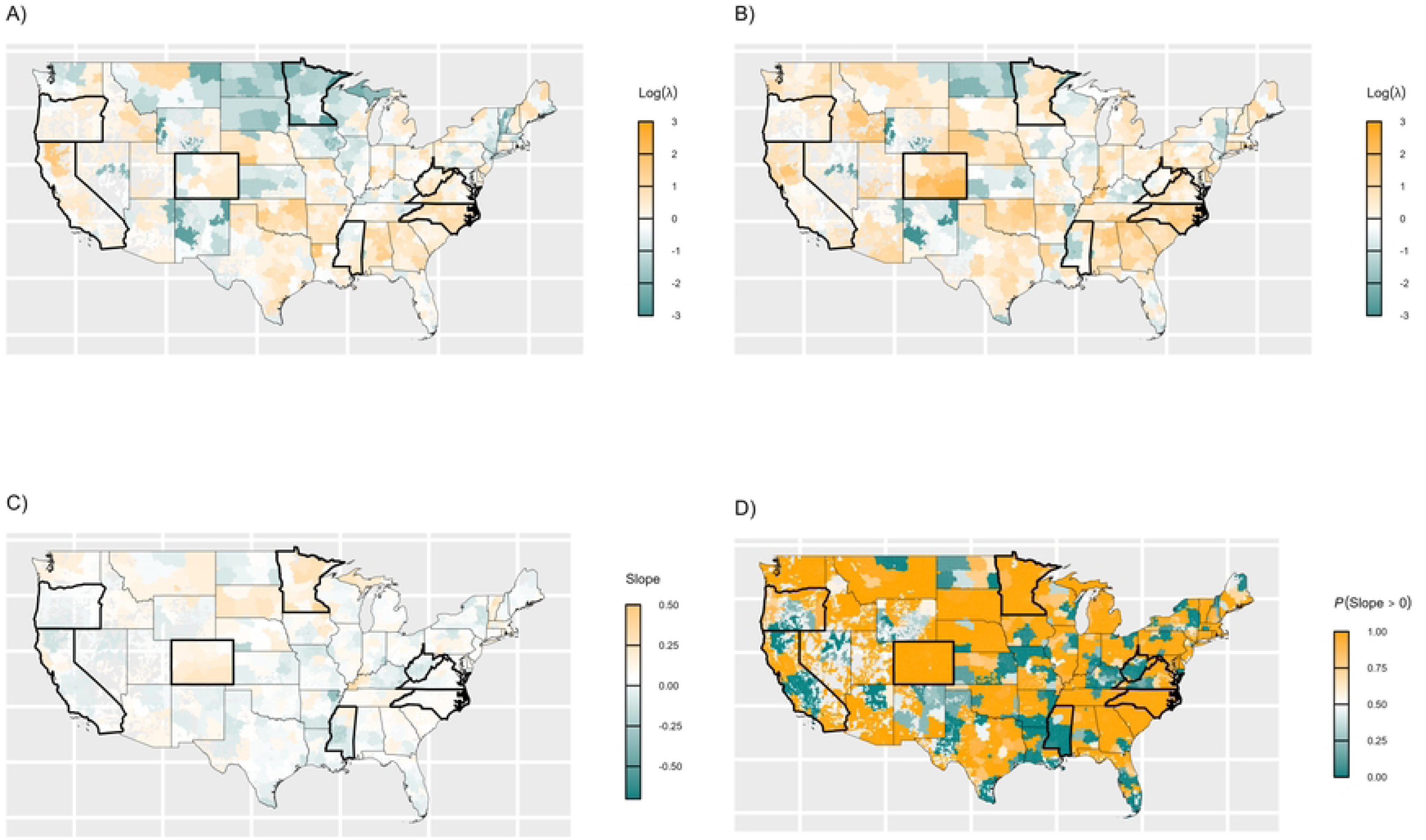
Geographic variation in pain-indicated buprenorphine prescribing, 2017-2025. Panels A and B show estimated prescribing rates relative to the 2017 national reference, displayed as log rate ratios, for three-digit ZIP code areas in 2017 and 2025. Panel C shows the estimated annual change on the log-rate scale; panel D shows the posterior probability that prescribing increased. Estimates come from the primary hierarchical model. Rate ratios are dimensionless. The eight illustrative states (Colorado, Minnesota, California, Oregon, Virginia, Mississippi, West Virginia, and North Carolina) are outlined in black.

### State Policy Approaches

The nationwide review identified clinical-guidance, coverage, oversight, and educational approaches with different timing and payer reach (Tables 1 and 2; Table B in S1 Appendix). Colorado combined earlier Medicaid opioid rules with a later commercial-coverage requirement; Minnesota had formulation-dependent prescribing-measure exclusions and distinct OUD and pain educational activities. Virginia paired an analgesic safety rationale with continued prior authorization. California and Oregon issued clinical guidance, while Connecticut and South Dakota removed specified coverage restrictions. Mississippi and West Virginia listed the buprenorphine patch (Butrans) as preferred in 2016, although Mississippi listed it as nonpreferred by 2018, and North Carolina preferred the patch under standard opioid criteria from 2018 without an enhanced pain-specific pathway. Table 2 reports dates, affected products, and scope, including provisions that preceded follow-up.

**Table 1.**
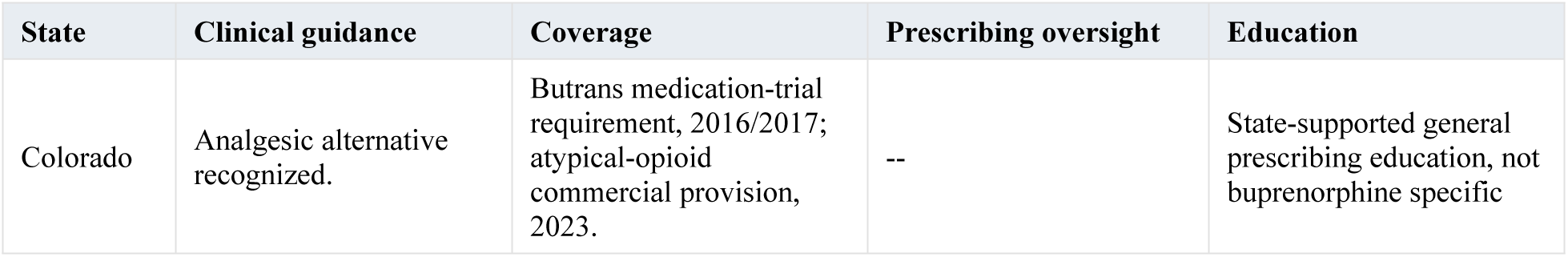
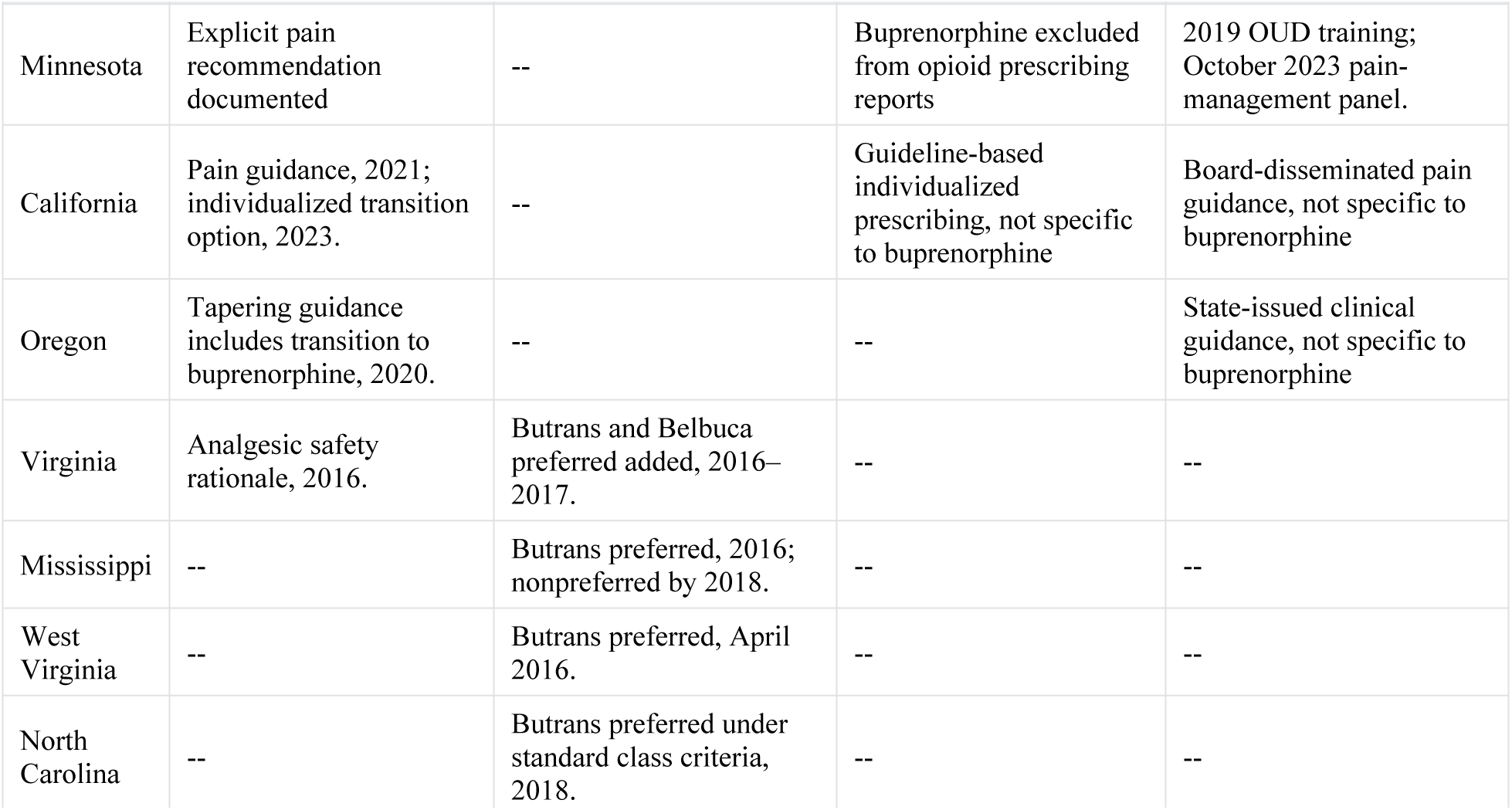
Selected State Policy Profiles from the Nationwide Review.

**Table 2.** Timing and Scope of Selected State Policy Actions.

| Date | State | Policy action |
|---|---|---|
| July 2016 and July 2017 | Colorado | Butrans listed with a single-step medication-trial requirement in both bulletins [21, 22]. |
| August 1 and October 1, 2017 | Colorado | Medicaid initial prescriptions limited to seven days; later MME threshold reduced from 300 to 250 mg/day [22]. |
| March 16 / November 15, 2018 | Colorado | Board opioid guidelines adopted March 16; Medicaid threshold reduced to 200 MME/day November 15. These were general opioid changes, not a verified patch-specific exemption [23, 24]. |
| June 28, 2021 / January 1, 2023 | Colorado | HB21-1276 signed / commercial coverage provisions effective. At least one pain-approved atypical opioid required at the lowest tier without PA or step therapy; no step therapy for additional atypical opioids. Not all buprenorphine products were guaranteed PA-free access [25]. |
| March / April 4, 2018 | Minnesota | Guideline document / public release. Technical prescribing-measure exclusions depended on formulation; a blanket pain-buprenorphine exemption was not established [26]. |
| February and December 2019 | Minnesota | Buprenorphine Boot Camps focused on OUD [27]. |
| October 17, 2023 | Minnesota | State work-group panel addressed pain management and buprenorphine [28]. |
| December 2025 | Minnesota | DHS report recommended promoting buprenorphine for chronic pain. Current DHS reporting excludes all buprenorphine products from prescriber comparisons used for quality improvement; the historical start of this broader exclusion is unverified [29, 30]. |
| 2016 / December 2016-April 2017 | Virginia | Butrans and Belbuca added as analgesic alternatives to Schedule II long-acting opioids based on lower respiratory risk; PA retained. FFS changes began July 2016; managed-care changes applied to new prescriptions December 2016 and all members April 2017 [31]. |
| January 2019 | Virginia | Medicaid expansion; separate from analgesic formulary policy [32]. |
| 2021 / July 2023 | California | Board pain-specific guidance / individualized transition guidance [33, 34]. |
| January 2020 | Oregon | Tapering guideline addressed transition to buprenorphine [35]. |
| 2018 | Arizona | Guideline advised considering buprenorphine rotation before tapering in selected patients; a conditional approach rather than a general maintenance recommendation [36]. |
| July 10, 2023 | Pennsylvania | Authorization criteria applied to FFS and managed Medicaid; PA and other opioid criteria remained [37]. |
| July 2016 / by June 2018 | Mississippi | Butrans preferred in July 2016 / listed as nonpreferred by June 2018; not a sustained preferred status [38, 39]. |
| April 2016 | West Virginia | Butrans listed as preferred [40]. |
| July 2018 | North Carolina | Butrans preferred under standard opioid criteria. Routine coverage was recorded, but no qualifying enhanced pain-specific pathway was identified under the review rubric [41–43]. |
| July 2021 / December 2023 | North Carolina | Managed-care launch / Medicaid expansion; separate from pain-formulation policy [44, 45]. |
| February 1, 2019 | Connecticut | Belbuca and Butrans removed from long-acting-opioid PA [46]. |
| October 14, 2019 | South Dakota | Belbuca and Butrans step therapy removed; date recorded in the state P&T historical summary [47]. |

OUD, opioid use disorder; PA, prior authorization. Profiles summarize distinct policy domains, with all 51 jurisdictions reported in Table B in S1 Appendix. Dashes denote no qualifying documented feature within the review scope. Routine preferred-drug placement under standard opioid criteria receives zero credit for enhanced access. Profiles are compared by content and timing; no composite score-gradient analysis was performed. Sources and dates appear in Table 2 and the cited policy documents. Minnesota’s exclusion means buprenorphine prescriptions do not contribute to the reported opioid measures. This could reduce concern about triggering quality-improvement review but is not a coverage exemption or evidence of increased prescribing.

DHS, Department of Human Services; FFS, fee-for-service; MME, morphine milligram equivalents; OUD, opioid use disorder; PA, prior authorization; P&T, Pharmacy and Therapeutics. Dates identify the event type. January 2018 is a nominated model break, not a policy-effective date.

### State Prescribing Growth in Relation to Policy Approaches

Growth varied widely among states with documented provisions (Table 3; Fig 2). Minnesota ranked first and Colorado fourth among 51 jurisdictions (25.43% and 13.50% annualized growth). North Carolina ranked 22nd (6.29%), exceeding national growth despite having routine preferred patch coverage without an enhanced pain-specific pathway. Virginia ranked 32nd (3.35%). Annualized growth was 2.99% in California, 0.63% in West Virginia, and 0.50% in Oregon; Mississippi declined 2.96%. Observed rates and rankings appear in Table 3 and Fig 2; Table K in S1 Appendix includes all jurisdictions.

**Fig 2.**
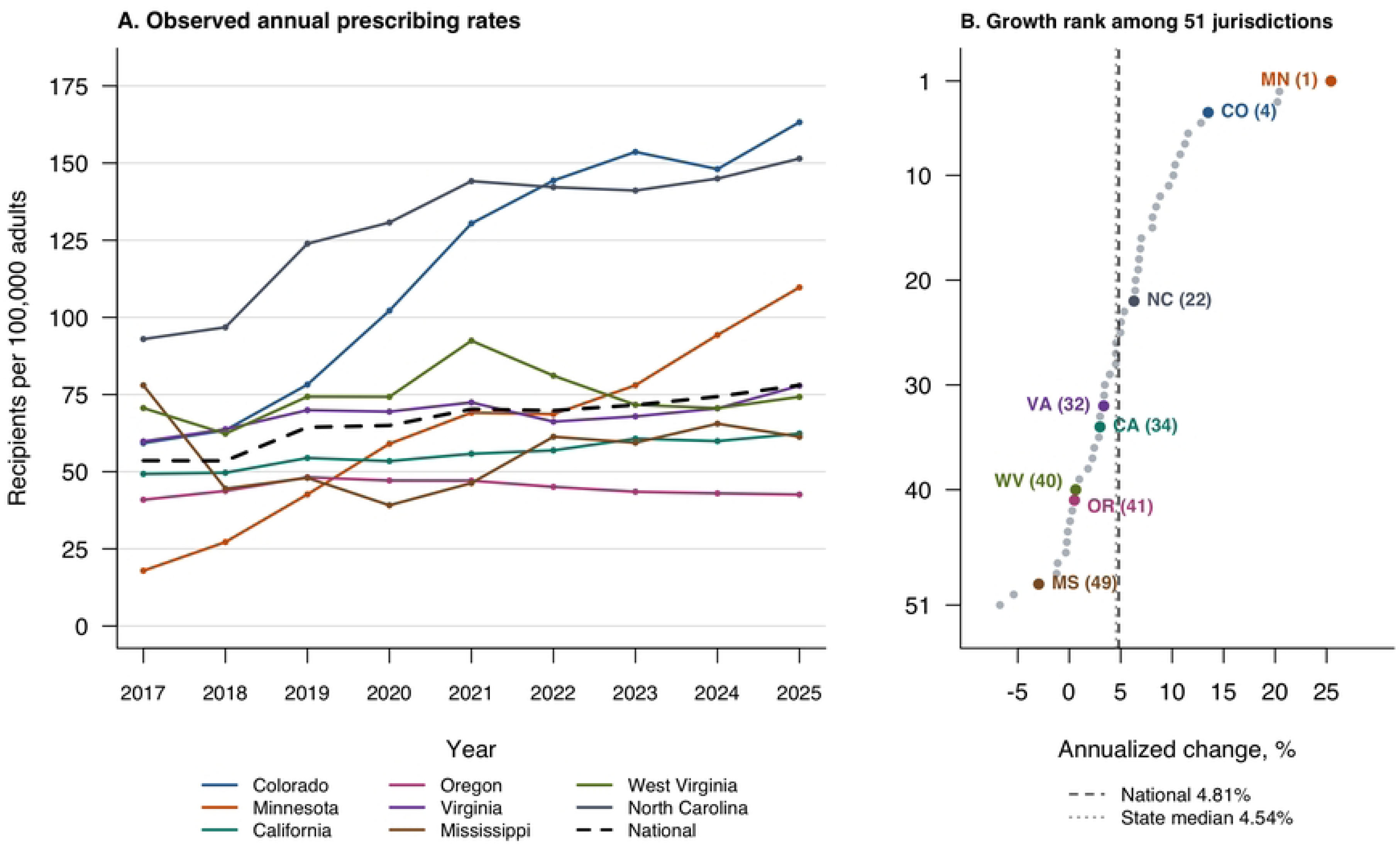
Observed national and selected-state prescribing rates and state growth rankings, 2017-2025. Panel A shows recipients per 100,000 adults using year-matched ACS five-year estimates; 2025 uses the 2024 denominator. The dashed black line is the population-weighted national comparator. Panel B ranks the 50 states and DC by annualized percentage change over eight years. Colorado, Minnesota, California, Oregon, Virginia, Mississippi, West Virginia, and North Carolina use matching colors in both panels. Colors identify states, not policy categories. Vertical lines mark national growth and the median jurisdiction. Connecting lines in Panel A are visual guides.

**Table 3.**
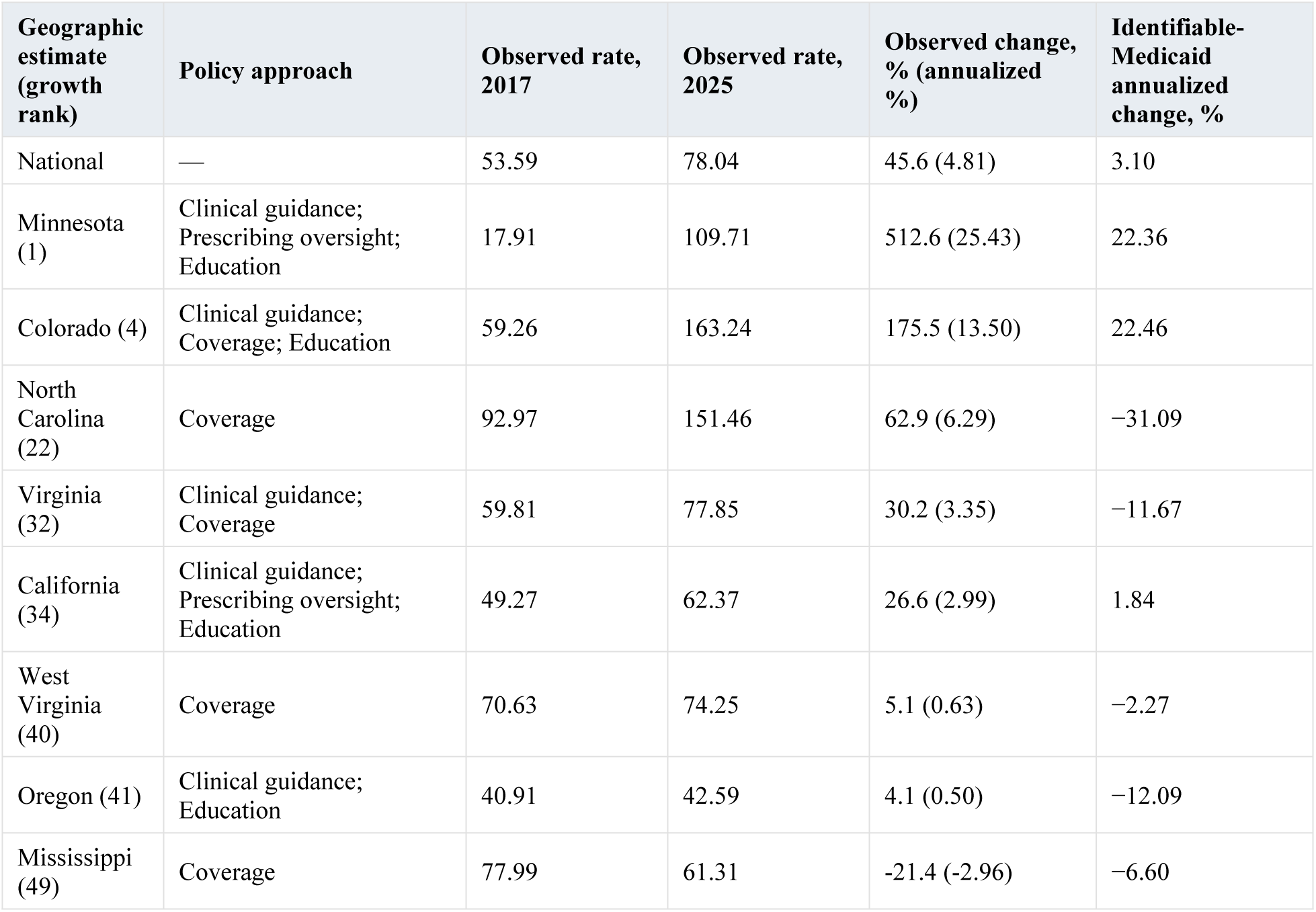
Observed Changes in Pain-Indicated Buprenorphine Prescribing, All Payers and Medicaid, 2017-2025.

| **Geographic estimate (growth rank)** | **Policy approach** | **Observed rate, 2017** | **Observed rate, 2025** | **Observed change, % (annualized %)** | **Identifiable-Medicaid annualized change, %** |
|---|---|---|---|---|---|
| National | — | 53.59 | 78.04 | 45.6 (4.81) | 3.10 |
| Minnesota (1) | Clinical guidance; Prescribing oversight; Education | 17.91 | 109.71 | 512.6 (25.43) | 22.36 |
| Colorado (4) | Clinical guidance; Coverage; Education | 59.26 | 163.24 | 175.5 (13.50) | 22.46 |
| North Carolina (22) | Coverage | 92.97 | 151.46 | 62.9 (6.29) | −31.09 |
| Virginia (32) | Clinical guidance; Coverage | 59.81 | 77.85 | 30.2 (3.35) | −11.67 |
| California (34) | Clinical guidance; Prescribing oversight; Education | 49.27 | 62.37 | 26.6 (2.99) | 1.84 |
| West Virginia (40) | Coverage | 70.63 | 74.25 | 5.1 (0.63) | −2.27 |
| Oregon (41) | Clinical guidance; Education | 40.91 | 42.59 | 4.1 (0.50) | −12.09 |
| Mississippi (49) | Coverage | 77.99 | 61.31 | -21.4 (-2.96) | −6.60 |

Observed rates use year-matched ACS five-year adult populations; 2025 uses 2024. Annualized change is compound change over eight years. All-jurisdiction observed results appear in Table K in S1 Appendix. Ranks compare annualized observed growth among all 50 states and DC; the national comparator is not ranked. Policy approaches use the four domains in Table 1; no state policy category is assigned to the national comparator. Model estimates are reported separately in the text and S1 Appendix. State rows are ordered by growth rank. Identifiable-Medicaid changes use the same adult population denominators (not Medicaid enrollment); identifiable Medicaid comprises fee-for-service and explicitly identified managed Medicaid plans, and plan coding and managed-care transitions can affect these trends. Payer-specific results for all jurisdictions appear in Table O in S1 Appendix.

Additional annual Bayesian trends were positive in Colorado, Minnesota, North Carolina, California, and Virginia, whereas intervals for Oregon, Mississippi, and West Virginia included zero (Table N in S1 Appendix). Dated policy comparisons varied by state and interval. Colorado’s rate rose 6.43% from 2022 to 2023, compared with 2.40% in the rest of the country. California’s 2020-2022 increase was 6.51%, compared with 7.51% elsewhere, and its 2022-2024 increase was 5.28%, compared with 6.57% (Table H and Fig E in S1 Appendix). In the monthly model, Colorado’s relative annual growth versus the rest of the country was 14.68% during 2018-2022 and 1.60% during 2023-2025, although estimates varied with model assumptions and nominated break dates (Table G and Fig C in S1 Appendix).

### Payer-Restricted Analyses

Payer-restricted results were not uniformly consistent with all-payer findings (Table 3). Nationally, identifiable-Medicaid recipients per 100,000 adults increased 3.10% annualized during 2017-2025 (fee-for-service only, 1.00%), compared with 4.81% for all payers. Identifiable-Medicaid annualized changes were 22.46% in Colorado and 22.36% in Minnesota, but −11.67% in Virginia and −31.09% in North Carolina, two states whose all-payer rates increased over the same period (Table O and Fig B in S1 Appendix). Among the remaining illustrative states, identifiable-Medicaid changes were 1.84% in California, −2.27% in West Virginia, −6.60% in Mississippi, and −12.09% in Oregon. These payer-specific estimates were sensitive to plan classification (Tables O and P in S1 Appendix). Mixed-plan claims that could not be assigned to Medicaid numbered 25,087 in 2017 and 47,839 in 2025, equivalent to 47%-57% of identifiable-Medicaid claims in each year (Table P in S1 Appendix). In 18 of 51 jurisdictions, the 2017 identifiable-Medicaid rate was below 1 per 100,000 adults, and payer-specific growth estimates for those jurisdictions are unstable (Table O in S1 Appendix).

### Sensitivity Analyses

Using a fixed population denominator or assigning each recipient to a single state yielded similar growth rankings; Minnesota and Colorado remained first and fourth, and rank correlations with the primary analysis were 0.992 and 0.999. Across Colorado, Minnesota, Virginia, and North Carolina, annualized estimates changed by no more than 1.45 percentage points with fixed 2024 denominators and 0.07 with dominant-state allocation (Table L in S1 Appendix). Geographic mapping completeness ranged from 97.3% to 99.3% (Table M in S1 Appendix). Detailed sensitivity findings appear in the S1 Appendix.

## DISCUSSION

Pain-indicated buprenorphine prescribing increased nationally from 2017 to 2025, supporting our hypothesis, but growth differed substantially across regions and states. A nationwide policy review identified several approaches that could influence access, yet states with documented policies had markedly different trajectories, and Medicaid-restricted patterns diverged from all-payer patterns in some of those states. The central finding is therefore a national increase accompanied by geographic heterogeneity that was not consistently related to the presence of a documented policy and that depended in part on which payers were examined.

This national increase in buprenorphine for pain occurred while overall opioid analgesic prescribing declined and buprenorphine initiation for OUD was relatively flat during much of the overlapping period [50, 51]. A separate study linked substantial state increases in OUD buprenorphine prescribing during 2018-2024 to increased Medicaid prescribing [52]. More directly relevant, Nguemeni Tiako and colleagues analyzed quarterly prescriptions among Medicaid enrollees, grouped by prior-authorization and preferred-drug status assessed in August 2025 [13]. Our study complements theirs by examining annual unique adult recipients rather than prescription volume, all-payer and payer-restricted rates per adult resident, ZIP3-level variation, and dated histories of policy, clinical guidance, and education during 2017-2025. The different numerators and denominators preclude direct numerical comparison of the rates.

National guidance provides context for these trends. The 2016 CDC guideline favored nonopioid therapy and preceded our observation period, so we could not assess a change from before its release [5]. The 2022 CDC guideline discussed transition to buprenorphine for selected patients, and the 2022 VA/DoD guideline suggested it over full-agonist opioids for patients receiving daily opioids for chronic pain [6, 53]. National rates rose after these two guidelines, from 69.82 per 100,000 adults in 2022 to 78.04 in 2025. However, the largest annual increase occurred earlier, in 2018-2019 (53.53 to 64.40; Table J in S1 Appendix). The later rise is temporally consistent with the guidance but does not establish its effect. National VA guidance further addressed pain management in 2024 [54]. Elimination of the federal X-waiver in 2023 changed OUD prescribing requirements; a waiver had never been required for pain prescribing, so its elimination would not be expected to affect pain-indicated prescribing directly [55]. Indirect effects through clinician familiarity, capacity, or stigma remain possible.

Against this national backdrop, the contrasting state trajectories suggest that policy content and implementation deserve attention beyond whether a written policy exists. For pain-indicated buprenorphine, Medicaid prescribing differed by prior-authorization and preferred-drug status, supporting attention to coverage restrictions rather than formulary listing alone [13]. A primary-care survey addressing chronic pain and OUD identified knowledge, confidence, cost, and insurance barriers, supporting evaluation of clinician education alongside coverage changes [56]. These studies and our findings do not establish that barrier reduction, education, or their combination caused greater uptake. Comparisons must also account for the starting prescribing rate, covered formulations, and payer reach: a policy introduced before 2017 may already have influenced the baseline, whereas a later Medicaid provision reaches only part of the population directly.

The payer-restricted analyses bear directly on how the state and national findings should be read. Many reviewed policies govern Medicaid coverage or oversight and can directly reach only Medicaid recipients, so all-payer estimates may dilute associations concentrated among those recipients. Cross-payer spillover is plausible. Medicaid expansion was associated with more buprenorphine prescribers serving Medicare Part D beneficiaries in an OUD study [57]. That finding does not establish spillover for pain treatment or a clinician-learning mechanism; neither was measured in our study. In Virginia and North Carolina, identifiable-Medicaid prescribing declined while all-payer prescribing rose, so all-payer growth in those states reflects non-Medicaid payers, Medicaid records that could not be identified as such, or both. North Carolina’s July 2021 managed-care transition, which moved most beneficiaries from fee-for-service into managed-care plans, further complicates payer-coded trends [44]. Because the national trend aggregates these state trajectories, the same payer composition shapes the national estimate: identifiable-Medicaid growth nationally (3.10% annualized) lagged all-payer growth (4.81%), and a population-based rate across all payers can rise while prescribing for the payer that state policies target changes little or declines. Medicaid expansion was a separate exposure, occurring in 2014 in Colorado and Minnesota, January 2019 in Virginia, and December 2023 in North Carolina [32, 45]. Although prior studies linked expansion to Medicaid-covered OUD treatment [58–60], our population-based rates lack enrollment denominators and do not isolate expansion effects.

For clinicians and policymakers, the geographic differences identify questions about whether guidance is clear, coverage is workable, and education addresses analgesia as well as OUD. Regional prescribing culture, pain burden, clinician supply, and insurance arrangements may also contribute. These findings support evaluating access alongside treatment appropriateness, pain relief, function, and safety; greater dispensing alone does not establish better care.

### Limitations

Claims identified formulations rather than diagnoses and did not establish treatment appropriateness, substitution for full-agonist opioids, or patient outcomes. Rates used adult population denominators, so geographic differences may partly reflect differences in chronic pain prevalence or in the number of adults receiving opioid analgesics rather than in the share of such patients receiving buprenorphine. Capture varied by payer and dispensing channel. The policy review classified documented policy content; implementation fidelity and individual exposure were unavailable.

Several policies preceded our follow-up time frame, and policy publication does not establish uptake. Our policy comparisons were exploratory, had relatively few post-event observations, and did not control all concurrent changes [61]. Nonsignificant estimates do not establish equivalence. Additional Bayesian intervals do not incorporate measurement error in claims, populations, or geographic allocation. Identifiable Medicaid is a lower bound: about one-third of Medicaid-like claims could not be assigned to Medicaid, capture varied widely across states, and if the unassigned plans include managed Medicaid, identifiable-Medicaid growth is understated wherever managed care expanded. Payer-restricted rates also lack enrollment denominators.

Annual prescribing trends and monthly models included 2025, using the 2024 population denominator for that year.

### Conclusions

Pain-indicated buprenorphine prescribing increased nationally with substantial state and regional variation. A nationwide policy review identified diverse approaches to clinical guidance, coverage, oversight, and education alongside widely differing prescribing trajectories, and Medicaid-restricted patterns did not consistently mirror all-payer trends. These findings support time-specific evaluation of how policy design, implementation, and payer reach influence access to buprenorphine for pain.

## Data Availability

Explanation for why data not available: The prescription claims data underlying this study were obtained under license from IQVIA (Longitudinal Prescription Claims database) and cannot be shared by the authors under the terms of the data use agreement. Interested researchers may license the same data directly from IQVIA (https://www.iqvia.com/insights/the-iqvia-institute/available-iqvia-data) or contact IQVIA at. The authors did not have any special access privileges to these data that other researchers would not have. Additional Information The complete nationwide policy review for all 50 states and the District of Columbia (Table B), the policy document inventory and classification rubric (Tables D and E), the pain-indicated formulation codes (Table C), and all aggregated results underlying the figures and tables (Tables F-Q and Figs A-E) are provided in S1 Appendix. The population denominators are publicly available from the American Community Survey (table B01001) and the HUD-USPS ZIP-County crosswalk. Analysis code for the hierarchical Bayesian model, the observed national and state rates, the payer-restricted analyses, the annual and monthly Bayesian log-rate trend models, and the policy comparisons is available from the corresponding author upon reasonable request.

## Supporting information

S1 Appendix. Supplementary methods, policy evidence, state results, sensitivity analyses, and completeness checks. Contains Tables A-Q and Figs A-E.

S2 Appendix. Data sharing statement.

